# Severity of COVID-19 at elevated exposure to perfluorinated alkylates

**DOI:** 10.1101/2020.10.22.20217562

**Authors:** P Grandjean, C.A.G. Timmermann, M. Kruse, F. Nielsen, P. Just Vinholt, L. Boding, C. Heilmann, K. Mølbak

## Abstract

**Background:** The course of coronavirus disease 2019 (COVID-19) seems to be aggravated by air pollution, and some industrial chemicals, such as the perfluorinated alkylate substances (PFASs), are immunotoxic and may contribute as well.

**Methods:** From Danish biobanks, we obtained plasma samples from 323 subjects aged 30-70 years with known SARS-CoV-2 infection. The PFAS concentrations measured at the background exposures included five PFASs known to be immunotoxic. Register data was obtained to classify disease status, other health information, and demographic variables. We used ordinal and ordered logistic regression analyses to determine associations between PFAS concentrations and disease outcome.

**Results:** Plasma-PFAS concentrations were higher in males, in subjects with Western European background, and tended to increase with age, but were not associated with the presence of chronic disease. Of the study population, 108 (33%) had not been hospitalized, and of those hospitalized, 53 (16%) had been in intensive care or were deceased. Among the five PFASs considered, perfluorobutanoic acid (PFBA) showed an odds ratio (OR) of 2.19 (95% confidence interval, CI, 1.39-3.46) for increasing severities of the disease, although the OR decreased to 1.77 (95% CI, 1.09, 2.87) after adjustment for age, sex, sampling site and interval between blood sampling and diagnosis.

**Conclusions:** Measures of individual exposures to immunotoxic PFASs included PFBA that accumulates in the lungs. Elevated plasma-PFBA concentrations were associated with an increased risk of more severe course of CIVID-19. Given the low background exposure levels in this study, the role of PFAS exposure in COVID-19 needs to be ascertained in populations with elevated exposures.

## Introduction

Elevated exposure to air pollution is associated with a worsened outcome of coronavirus disease 2019 (COVID-19).^1-4^ While replicated in different populations, the evidence relies on ecological study designs without measures of individual levels of exposure. Several industrial chemicals can suppress immune functions^5,6^ and worsen the course of infections.^7^ In particular, the perfluorinated alkylate substances (PFASs) are persistent, globally disseminated chemicals known to be immunotoxic.^8^ Thus, elevated PFAS exposure is associated with lower antibody responses to vaccinations in children^9^ and in adults.^10^ Also, infectious disease occurs more frequently in children with elevated background exposure.^11-13^ Further, major PFASs are suspected interfering with proteins involved in critical pathways associated with severe clinical outcomes of the COVID-19 infection.^14^

Substantial differences occur in the clinical course of the disease, and the reasons for this variability are only partially known.^15,16^ As a possible contributor, weak specific antibody responses may be an important contributor to a more severe clinical course of the infection,^17^ as also suggested by the poorer prognosis in patients with bacterial co-infection.^18^ The most serious clinical consequences are associated with male sex, older age, and the presence of co-morbidities, including obesity and diabetes.^19-23^ In parallel, serum-PFAS concentrations are higher in men than in women and also tend to increase with age.^8,24^ Because PFAS exposure has been linked to both obesity and diabetes,^25,26^ PFAS exposure may potentially affect the progression of COVID-19 directly as well as indirectly.

Several PFASs can be reliably determined in human blood samples, where they show a long biological half-life of 2-3 years or more,^27^ thereby providing a measure of cumulated exposure. Still, blood concentrations may not accurately reflect the retention in specific organs, e.g., the short-chain perfluorobutanoic acid (PFBA), which accumulates in the lungs.^28^

To assess if elevated background exposures to immunotoxic PFASs are associated with the clinical course of the infection, a study was undertaken in Denmark to determine individual plasma-PFAS concentrations in adults confirmed to be infected with severe acute respiratory syndrome coronavirus 2 (SARS-CoV-2) and examine the association with the severity of COVID-19 development.

## Methods

### Population

Plasma samples for PFAS analysis were obtained from medical biobanks that store excess material from diagnostic tests, viz., the Danish National Biobank at the Statens Serum Institut (SSI) and Odense University Hospital (OUH). Eligible subjects were identified from the Danish cohort of COVID-19 patients.^29^ All cases were tested by quantitative polymerase-chain-reaction (PCR) and had a positive response for SARS-CoV-2 infection, as recorded in the Danish Microbiology Database (MiBa), a national database that contains both positive and negative results of the majority of microbiology testing done in Denmark.^30^

The study included non-pregnant subjects aged 30-70 years at the time of the positive test by early March 2020 through early May 2020, provided that the biobanks could provide a plasma sample of 0.15 mL. Although most blood samples were obtained soon after SARS-CoV-2 infection was identified, we also included subjects, mainly those not hospitalized, whose plasma in the SSI biobank had been obtained up to 28 months earlier, i.e., less than a half-life for major PFASs.^27^

All samples were coded, and the Personal Identification Number for each subject was separately transferred to the Danish Health Data Authority (FSEID-00005000) for linkage to demographic and medical information from the Danish Civil Registration System (CRS),^31^ the Danish National Register of Patients (DNRP),^32^ and the National Health Insurance Service Register.^33^ By linkage to the national patient register, we classified disease status as follows: no hospital admission and completed infection within 14 days of testing positive, hospitalization with COVID-19 up to or above 14 days, admission to intensive care unit, or death. The linked data set was analyzed via secure server without access to information on the identity of the subjects involved. For confidentiality reasons, all tabular information was based on no less than five subjects.

The protocol was approved by the Regional Committee on Health Research Ethics (S-20200064), which also allowed the project to proceed without seeking informed consent from the subjects identified for study participation. Additional approvals were obtained from the Danish Data Protection Agency as well as institutional and regional authorities for the transfer blood samples and linkage of subject information to the PFAS analyses, while protecting confidentiality.

### Chemical Analysis

The plasma samples were analyzed in succeeding series for PFAS concentrations, including PFBA, perfluorooctane sulfonate (PFOS), perfluorooctanoate (PFOA), perfluorohexane sulfonate (PFHxS), and perfluoronanoate (PFNA). We used online solid-phase extraction followed by liquid chromatography and triple quadropole mass spectrometry (LC–MS/MS) at the University of Southern Denmark.^34^ Accuracy of the analysis was ensured by inclusion of quality control (QC) samples comprising proficiency test specimens from the HBM4EU program organized by Interlaboratory Comparison Investigations (ICI) and External Quality Assurance Schemes (EQUAS). All results of the QC samples were within the acceptance range. The between-batch CVs for the actual series ranged between 3% and 14% for all compounds. Both PFOS and PFOA were quantified in all blood samples, and all PFASs were detectable in at least 30% of the samples. Results below the limit of detection (LOD, 0.03 ng/ml) were replaced by LOD/2 before uploading to the secure server at the Danish Health Data Authority, where linkage to other information took place.

### Statistical Analysis

Correlation between PFASs were examined using Spearman’s correlation coefficient. The PFAS concentrations were compared between demographic groups and presence of comorbidities and tested using Kruskal-Wallis test of variance and Wilcoxon rank-sum test. Furthermore, associations between demographic groups and COVID-19 severity were tested using χ^2^ test. Associations between comorbidities, place of inclusion, and COVID-19 severity could not be displayed and tested due to confidentiality concerns, as some cells contained less than five individuals.

The association between plasma-PFAS concentrations and COVID-19 severity were tested in ordered logistic regression models. Because more than half the PFBA and PFBS concentrations were below the LOD, they were included as binary variables (below/above LOD). Potential confounding variables were identified based on a priori knowledge and included age (continuous, years) sex, and national origin (Western European yes/no). Among those of Western European national origin, 94% were Danish, while subjects born in or of parents from Somalia (20% of the group), Pakistan (13%), Iraq (12%), Morocco (11%), Eastern Europe (9%), and Turkey (9%) constituted most of the participants of non-Western European national origin. Because past PFAS exposure could potentially increase the risk of certain chronic diseases that may affect COVID-19 severity, chronic disease (yes/no) was considered a potential confounder to allow estimation of the direct, rather than the total effect of PFAS exposure. Place of inclusion (OUH/SSI) and timing of blood sampling were associated with both PFAS concentrations and COVID-19 severity and were therefore included as covariates.

The assumption of PFAS linearity was tested by including PFAS squared along with PFAS in the regression models. No significant (p<0.05) deviation from linearity was found. The proportional odds assumption in the ordinal logistic regression was tested using a likelihood-ratio test using the Stata *omodel* package. The test could not be fitted on the full model, but in a model adjusting for age, place of inclusion, and timing of blood sampling, the hypothesis of proportional odds was accepted (p>0.05) in all analyses. Odds ratios (ORs) between groups of COVID-19 severity were calculated using logistic regression models.

## Results

The predominant PFAS in plasma was PFOS, with an average concentration of 6.1 ng/mL (median, 4.7 ng/L), approximately equally distributed between the normal and branched isomers. Other PFASs quantified showed averages below 1 ng/mL. In a sensitivity analysis, one extreme PFHxS outlier at 12.9 ng/mL was omitted. The PFAS concentrations correlated well, with Spearman correlation coefficients generally above 0.5 (Table 1), except for PFBA. PFOS on average contributed 69% of the total PFAS concentrations and correlated particularly well with the most other PFASs quantified.

**Table 1.** Spearman’s correlation coefficients for pairwise comparisons of detectable PFASs in plasma from 323 subjects included in the study

|  | **PFBA** | **PFHxS** | **PFOS** | **PFOA** |
| --- | --- | --- | --- | --- |
| **PFHxS** | 0.0520 |  |  |  |
| **PFOS** | 0.0591 | 0.8406 |  |  |
| **PFOA** | 0.0617 | 0.7072 | 0.7248 |  |
| **PFNA** | 0.0127 | 0.7133 | 0.8406 | 0.7759 |

In general, serum-PFAS concentrations were higher at older ages, in men, and among those of Western European origin, though not in the presence of chronic disease (Table 2). PFBA was lower, but the origin of the samples was only weakly associated with plasma-PFAS concentrations (Table 2).

**Table 2.** Median plasma-PFAS concentrations (25^th^, 75^th^ percentiles) in ng/mL by population characteristics

| **Population characteristics** | **No. of persons (%)** | **PFBA** | **PFHxS** | **PFOS** | **PFOA** | **PFNA** |
| --- | --- | --- | --- | --- | --- | --- |
| **Total** | 323 (100) | <LOD (<LOD, 0.04) | 0.48 (0.28, 0.71) | 4.86 (2.85, 8.29) | 0.77 (0.43, 1.18) | 0.38 (0.23, 0.59) |
| **Age (years)** |  |  |  |  |  |  |
| 30-39 | 37 (11) | <LOD (<LOD, 0.03) | 0.32 (0.19, 0.46) | 3.30 (1.89, 5.27) | 0.59 (0.43, 0.86) | 0.29 (0.21, 0.43) |
| 40-49 | 64 (20) | <LOD (<LOD, 0.03) | 0.35 (0.15, 0.57) | 3.11 (2.24, 5.06) | 0.58 (0.35, 0.89) | 0.27 (0.19, 0.39) |
| 50-59 | 106 (33) | <LOD (<LOD, <LOD) | 0.50 (0.31, 0.75) | 5.41 (2.79, 8.84) | 0.83 (0.43, 1.18) | 0.40 (0.24, 0.61) |
| 60-70 | 116 (36) | <LOD (<LOD, 0.05) | 0.56 (0.39, 0.89) | 6.11 (3.83, 9.60) | 0.97 (0.56, 1.51) | 0.48 (0.30, 0.70) |
| *P* value <sup>a</sup> |  | 0.008 | <0.001 | <0.001 | <0.001 | <0.001 |
| **Sex** |  |  |  |  |  |  |
| Male | 174 (54) | <LOD (<LOD, 0.04) | 0.59 (0.40, 0.87) | 5.96 (3.65, 10.17) | 0.81 (0.51, 1.26) | 0.40 (0.25, 0.61) |
| Female | 149 (46) | <LOD (<LOD, 0.04) | 0.35 (0.17, 0.52) | 3.43 (2.06, 5.66) | 0.70 (0.40, 1.04) | 0.36 (0.22, 0.56) |
| *P* value <sup>b</sup> |  | 0.713 | <0.001 | <0.001 | 0.011 | 0.131 |
| **Chronic disease** |  |  |  |  |  |  |
| Yes | 222 (69) | <LOD (<LOD, 0.04) | 0.46 (0.28, 0.67) | 4.66 (2.88, 7.87) | 0.71 (0.42, 1.15) | 0.37 (0.23, 0.57) |
| No | 101 (31) | <LOD (<LOD, 0.03) | 0.51 (0.28, 0.76) | 5.35 (2.72, 8.41) | 0.87 (0.47, 1.20) | 0.42 (0.23, 0.65) |
| *P* value <sup>b</sup> |  | 0.069 | 0.262 | 0.726 | 0.147 | 0.309 |

| National origin |  |  |  |  |  |  |
| --- | --- | --- | --- | --- | --- | --- |
| Western Europe | 224 (69) | <LOD (<LOD,<br>0.04) | 0.52 (0.35,<br>0.76) | 5.61 (3.40,<br>9.18) | 0.91 (0.60,<br>1.29) | 0.43 (0.29,<br>0.64) |
| Other | 99 (31) | <LOD (<LOD,<br>0.04) | 0.34 (0.16,<br>0.57) | 2.86 (1.61,<br>5.13) | 0.44 (0.31,<br>0.80) | 0.23 (0.16,<br>0.36) |
| *P* value <sup>b</sup> |  | 0.552 | <0.001 | <0.001 | <0.001 | <0.001 |
| Place of inclusion |  |  |  |  |  |  |
| Odense | 48 (15) | <LOD (<LOD,<br>0.06) | 0.45 (0.32,<br>0.69) | 4.67 (3.29,<br>8.09) | 0.67 (0.42,<br>0.95) | 0.36 (0.24,<br>0.45) |
| Copenhagen area | 275 (85) | <LOD (<LOD,<br>0.03) | 0.48 (0.28,<br>0.72) | 4.89 (2.72,<br>8.31) | 0.79 (0.44,<br>1.20) | 0.39 (0.23,<br>0.62) |
| *P* value <sup>b</sup> |  | 0.003 | 0.967 | 0.697 | 0.203 | 0.299 |
| Timing of blood sampling |  |  |  |  |  |  |
| After diagnosis - 1<br>month before | 199 (62) | <LOD (<LOD,<br>0.04) | 0.48 (0.30,<br>0.71) | 4.63 (2.83,<br>7.65) | 0.70 (0.40,<br>1.11) | 0.35 (0.23,<br>0.56) |
| >1 month - 1 year<br>before diagnosis | 40 (12) | <LOD (<LOD,<br>0.02) | 0.43 (0.22,<br>0.70) | 5.06 (2.06,<br>8.76) | 0.85 (0.38,<br>1.35) | 0.37 (0.21,<br>0.67) |
| > 1 year before<br>diagnosis | 84 (26) | <LOD (<LOD,<br>0.03) | 0.50 (0.30,<br>0.72) | 5.48 (3.10,<br>10.28) | 0.87 (0.57,<br>1.22) | 0.45 (0.28,<br>0.65) |
| *P* value <sup>a</sup> |  | 0.182 | 0.819 | 0.204 | 0.074 | 0.050 |

<sup>a</sup> Variables with more than two categories tested using Kruskal-Wallis rank test
<sup>b</sup> Binary variables tested using Wilcoxon rank-sum test

In the study population, males, older subjects, and those with chronic disease, were more frequently represented among subjects with severe COVID-19, while there was no difference in regard to national origin for disease severity (Table 3).

**Table 3.**
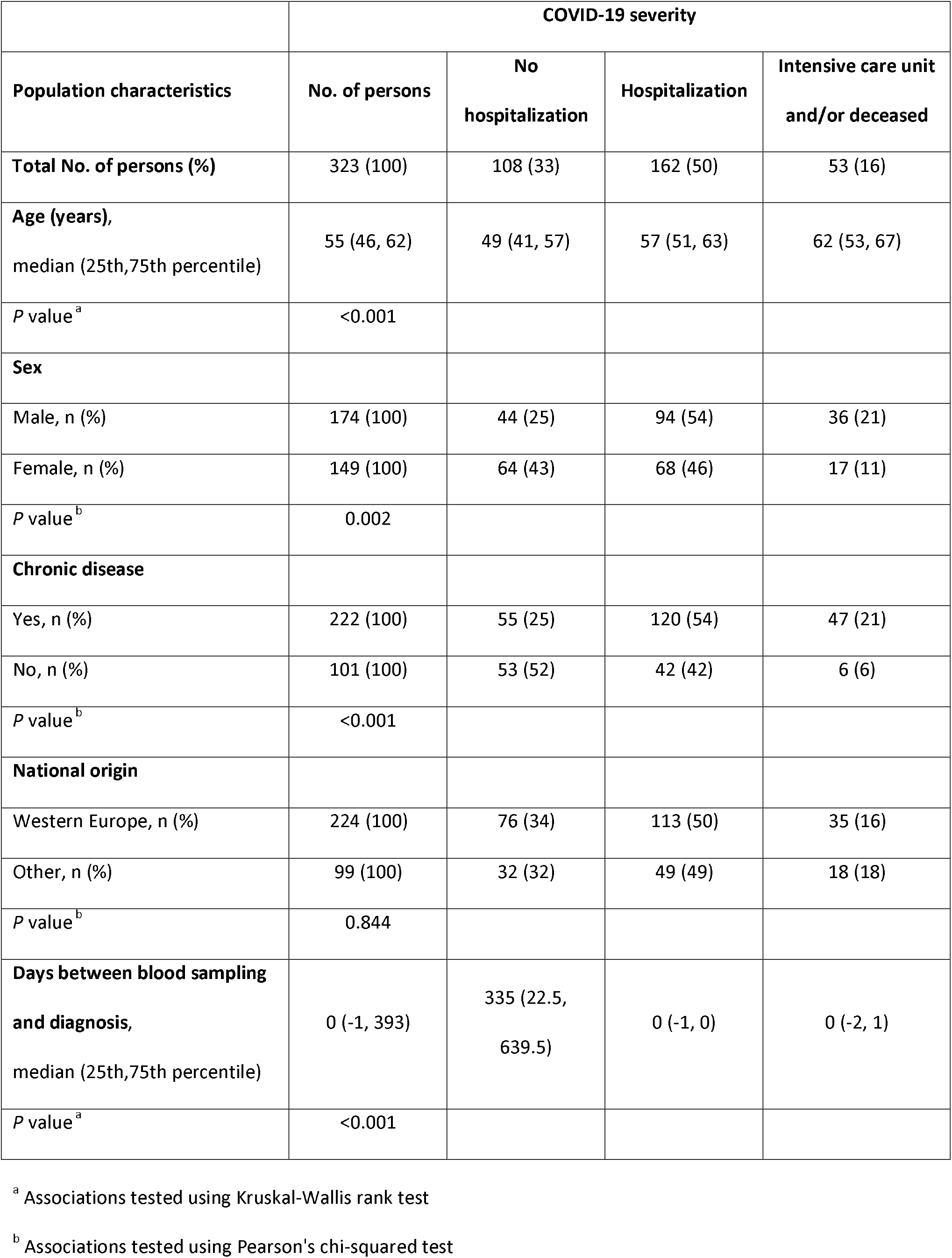
COVID-19 severity by population characteristics

A more severe disease outcome was associated with higher plasma-PFBA concentrations, also after adjustment for all covariates (Table 4). None of the other PFASs showed such tendency, and some were associated with lower risk. The PFAS-associations with disease severity were similar in Western Europeans and subjects with other backgrounds (*P* > 0.2 for population differences). In additional analyses, marginal changes in the ORs occurred when plasma samples obtained more than 60 days before diagnosis were excluded. If not adjusted for the presence of chronic disease, the adjusted OR for PFBA was 1.77 (95% CI, 1.09, 2.87); other ORs also changed only marginally.

**Table 4.** Ordered logistic regression OR of increased Covid-19 severity for an increase by 1 ng/mL in plasma-PFAS concentrations

| **PFAS** | **No. of persons** | **Crude OR (95% CI)** | **Adjusted OR (95% CI)<sup>a</sup>** | **Western European origin OR (95% CI)<sup>b</sup>** | **Non-western European origin OR (95% CI)<sup>b</sup>** |
| --- | --- | --- | --- | --- | --- |
| PFBA<br>(>LOD/<LOD) | 104/219 | 2.19 (1.39, 3.46) | 1.62 (0.99, 2.64) | 1.50 (0.83, 2.71) | 1.89 (0.80, 4.49) |
| PFHxS (ng/mL) | 323 | 0.85 (0.63, 1.15) | 0.52 (0.29, 0.91) | 0.46 (0.23, 0.89) | 0.70 (0.25, 1.93) |
| PFHxS<sup>c</sup> (ng/mL) | 322 | 1.00 (0.62, 1.61) | 0.52 (0.29, 0.93) | 0.46 (0.23, 0.90) | 0.70 (0.25, 1.93) |
| PFOS (ng/mL) | 323 | 1.00 (0.96, 1.04) | 0.97 (0.92, 1.02) | 0.97 (0.92, 1.03) | 0.98 (0.87, 1.10) |
| PFOA (ng/mL) | 323 | 0.99 (0.72, 1.36) | 0.87 (0.60, 1.26) | 0.77 (0.50, 1.18) | 1.25 (0.61, 2.52) |
| PFNA (ng/mL) | 323 | 1.18 (0.67, 2.09) | 1.11 (0.58, 2.13) | 0.97 (0.44, 2.13) | 1.52 (0.48, 4.78) |

<sup>a</sup> Adjusted for age (years), sex, chronic disease (yes/no), national origin (western European/non-western European), place of testing (Odense/Copenhagen area), and days between blood sampling and diagnosis
<sup>b</sup> Adjusted for age (years), sex, chronic disease (yes/no), place of testing (Odense/Copenhagen area), and days between blood sampling and diagnosis
<sup>c</sup> PFHxS >10 ng/mL excluded

In dichotomous analyses comparing severities of the disease, detectable PFBA in plasma also showed a clear association with a more severe clinical course of the disease, most pronounced for odds between hospitalization and admission to intensive care unit/death (data not shown). The association between PFBA and severity was similar for men and women (Figure 1).

**Figure 1.**
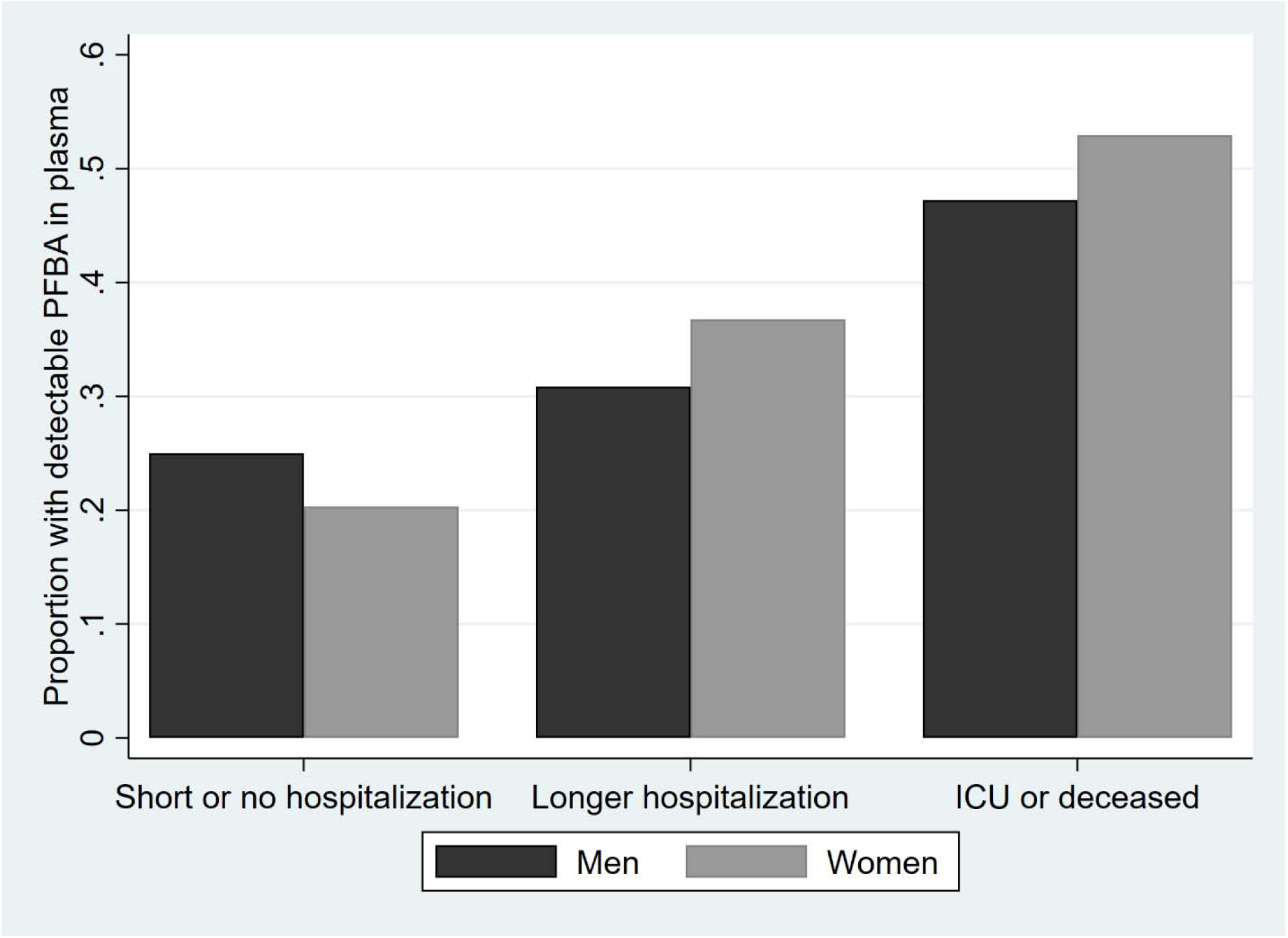
Proportion of plasma samples with detectable PFBA concentrations at different disease severities. Results are shown for 44 men and 64 women with up to two weeks of hospitalization, 94 men and 68 women with longer hospitalization, and 36 men and 17 women admitted to the intensive care unit (ICU) or deceased (*P* = 0.003).

## Discussion

The present study aimed at determining the potential aggravation of COVID-19 associated with elevated exposures to PFASs. Several of these substances are known immunotoxicants in laboratory animals^35^ and in humans.^8,9^ In addition to immunotoxicity, major PFASs can potentially interfere with major pathways that are predictive of a serious clinical outcome of the infection.^14^ An association of PFAS exposure with disease severity therefore appears biologically plausible.

Among the PFASs, presence of detectable PFBA in plasma showed the strongest association with the severity of the disease. This finding may at first seem surprising, as this PFAS has a short elimination half-life in the blood and is often considered of less importance to health.^27^ However, in tissue samples from autopsies, PFBA is the only PFAS that is substantially accumulated in the lungs.^28^ Given the persistence of the PFASs in general, the unique retention of PFBA in lung tissue may offer a clue to interpreting the findings in this study.

The associations of PFASs with a more serious course of COVID-19 are weakened after adjustment for covariates, and some regression coefficients and ORs are below 1. However, adjustment for all covariates may result in over-adjustment bias. Thus, older age and male sex are known to be strong predictors of higher blood-PFAS concentrations, and simple adjustment for these factors could potentially result in a bias toward the null. As PFAS exposure has been linked to important comorbidities, such as diabetes and obesity,^25,26^ both of which may exacerbate the virus infection, adjustment for chronic disease may also not be justified. Leaving it out slightly strengthened the PFBA association with the disease severity. Also, in consideration of the low background exposure levels, the fully adjusted results should therefore not be interpreted as evidence that most PFASs do not contribute to a worsened clinical course of COVID-19.

The results of this study are parallel to findings in regard to other environmental toxicants, viz., air pollutants^1-4^ and suggests a need to ascertain the impact of relevant occupational or environmental exposures on COVID-19 severity. Of note, the evidence on air pollution relies solely on ecological study designs without measures of individual levels of exposure, while the present study benefitted from measurements of plasma-PFAS concentrations of all study subjects.

In regard to limitations, the study population is not representative of corona-positive subjects, as inclusion in the study depended solely on the existence of plasma from diagnostic blood samples at the hospitals. Thus, subjects with chronic disease or more severe COVID-19 likely had more frequent hospital visits or longer admissions and thereby a greater chance of having plasma available for inclusion in this study. With a corona-related fatality rate of Danish blood donors below 70 years of age at 89 per 100,000 infections,^36^ the presence of 17 deaths in the present material (i.e., against 0.3 deaths expected) confirms that the blood samples represent a highly selected population. Still, a total of 108 subjects were known to have been infected, though not hospitalized. In many cases, their plasma had been stored on previous occasions, and the PFAS concentrations may reflect slightly higher exposures in the recent past.^8^ Although adjustment for the time interval since sample collection was included in the analyses, its impact on the results was negligible.

The study population included relatively older subjects who were more frequently male, and a large proportion of foreign-born subjects and second-generation immigrants (Table 3), thereby possibly also deviating from the background population of corona-infected patients in Denmark. Still, such biases will not necessarily affect the PFAS exposure and its association with COVID-19 outcomes.

Among immigrants, adverse associations appeared slightly stronger, also after adjustments, thus suggesting that national origin, perhaps as related to demographic or social factors, may result in a greater vulnerability to PFAS-associated aggravation of the infection. Difference in age, sex, or comorbidities did not explain this tendency, but is in agreement with previous findings of ethnic differences in vulnerability.^37^ However, national origin may be a surrogate marker for other factors, such as exposure at work or exposure within crowded households, as immigrant origin tends to be associated with certain occupations including front-line workers and living in areas with higher population density.^38^

## Conclusions

Increased plasma-PFBA concentrations were associated with a greater severity of COVID-19 prognosis, and this tendency remained after adjustment for sex, age, comorbidities, national origin, sampling location and time. Although PFBA occurred in lower plasma concentrations than most other PFASs determined, it is known to accumulate in the lungs. Thus, given the immunotoxicity of the PFASs, exposure to these persistent industrial chemicals may contribute to the severity of COVID-19. These findings at background exposure levels suggest a need to ascertain if exposures to environmental immunotoxicants may worsen the outcome of the SARS-CoV-2 infection.

## Data Availability

Data contain personal information on study participants and were uploaded to the secure server at the Danish Health Data Authority, where they are available pending necessary approvals. Aggregated data that involve at least five subjects are available from the authors.

## Acknowledgments

The Danish Departments of Clinical Microbiology, the Danish Microbiology Database and the section for Data Integration and Analysis at Statens Serum Institut collated the national COVID-19 data. Plasma samples were identified and provided by Statens Serum Institut and the Department of Clinical Biochemistry and Pharmacology at Odense University Hospital.

## Author Contributions

Conceptualization: Philippe Grandjean, Amalie Timmermann, Marie Kruse, Lasse Boding, Carsten Heilmann, Kåre Mølbak.

Data curation: Philippe Grandjean, Amalie Timmermann, Marie Kruse, Pernille Vinholt, Flemming Nielsen, Lasse Boding.

Formal analysis: Philippe Grandjean, Amalie Timmermann, Marie Kruse.

Funding acquisition: Philippe Grandjean, Lasse Boding, Kåre Mølbak.

Investigation: Philippe Grandjean, Amalie Timmermann, Marie Kruse, Pernille Vinholt, Flemming Nielsen, Lasse Boding.

Methodology: Philippe Grandjean, Amalie Timmermann, Marie Kruse, Pernille Vinholt, Flemming Nielsen, Lasse Boding.

Project administration: Philippe Grandjean, Kåre Mølbak.

Resources: Philippe Grandjean, Pernille Vinholt, Flemming Nielsen, Lasse Boding, Kåre Mølbak.

Software: Amalie Timmermann, Marie Kruse.

Supervision: Philippe Grandjean, Kåre Mølbak.

Validation: Philippe Grandjean, Amalie Timmermann, Marie Kruse, Lasse Boding.

Visualization: Marie Kruse.

Writing – original draft: Philippe Grandjean.

Writing – review & editing: Philippe Grandjean, Amalie Timmermann, Marie Kruse, Pernille Vinholt, Flemming Nielsen, Lasse Boding, Carsten Heilmann, Kåre Mølbak.

